# Determining the period of communicability of SARS-CoV-2: A rapid review of the literature

**DOI:** 10.1101/2020.07.28.20163873

**Authors:** Mina Park, Colleen Pawliuk, Tribesty Nguyen, Amanda Griffitt, Linda Dix-Cooper, Nadia Fourik, Martin Dawes

## Abstract

**Introduction:** How long individuals may transmit virus after infection with severe acute respiratory syndrome coronavirus 2 (SARS-CoV-2) is unclear. Understanding the communicability period of SARS-CoV-2 is important to inform the period of isolation required to prevent nosocomial and community spread. The objective of this study was to identify the reported communicable period of SARS-CoV-2, based on a rapid review of existing literature.

**Methods:** Studies reporting empirical data on the period of communicability of SARS-CoV-2 through investigations of duration of communicability based on in-person contact (“contact transmission”), isolation and culture of virus (“viral isolation”), and viral shedding by detection of nucleic acids by RT-PCR (“viral shedding”) were identified through searches of peer-reviewed and pre-print health sciences literature databases (Ovid MEDLINE, Embase, Google Scholar, medRxiv and arXiv) and the grey literature. Articles were screened for relevance, then data were extracted, analyzed, and synthesized.

**Results:** Out of the 165 studies included for qualitative analysis, one study investigated contact transmission, three investigated viral isolation, 144 investigated viral shedding, and 17 studies focused on both viral shedding and viral isolation. The median length of time until viral clearance across all viral isolation studies was nine days; however, the maximum identified duration was 32 days. Studies with data on both viral isolation and viral shedding showed a prolonged maximum time until viral clearance for viral shedding (9 days vs 24 days).

**Discussion:** Findings from this review support a minimum 10-day period of isolation; however, additional observation should be considered for individuals being released into high-risk settings.

## Introduction

Understanding how long individuals may continue to transmit virus after infection with severe acute respiratory syndrome coronavirus 2 (SARS-CoV-2) is important to inform policies on the period of isolation required to prevent nosocomial and community spread. There are two main strategies on discontinuation of isolation for cases [1–3]. The first, a test-based strategy, relies on resolution of symptoms accompanied by single or consecutive negative results from reverse transcriptase polymerase chain reaction (RT-PCR)-based tests, taken from respiratory samples. The second approach requires adherence to a set period of isolation (typically, 10 - 14 days from symptom onset or from the first positive test) in addition to resolution of symptoms.

While a test-based strategy may be preferred as providing an indication of viral clearance, this approach has important limitations. Viral shedding, as detected by the presence of viral nucleic acid by RT-PCR, does not equate to infectiousness as nucleic acid tests do not differentiate between live (or viable) and non-infective virus [4]. Moreover, the performance of SARS-CoV-2 PCR-based tests is inconsistent, with inconclusive test results being reported based on variations in individual viral shedding patterns. Anecdotal reports have emerged of individuals remaining in prolonged isolation due to continuous positive RT-PCR results, long after resolution of symptoms, with unintended adverse consequences [5]. Due to these issues, a fixed quarantine period and symptom resolution continues to be an important alternative or complimentary strategy.

To inform the fixed quarantine period strategy, it is important to understand as much as possible about the infectious period (called the communicability period in the rest of this paper). As such, the goal of this study was to identify reported communicable periods of SARS-CoV-2, based on a comprehensive review of existing literature. From an initial investigation into this topic, previous reviews were identified, a rapid scoping review and a systematic review, that investigated the potential duration of communicability [6,7]. However, a preliminary assessment of results from an independent search identified a number of additional studies that were not captured in either of these reviews. As these subsequent studies may alter the previous review findings, this review was conducted to include a greater number of studies, and more recent ones, than previous reports. Moreover, the scope of this review was specifically intended to include studies reporting empirical data on the period of communicability of SARS-CoV-2 through investigations of duration of communicability based on in-person contact (“contact transmission”), isolation and culture of virus (“viral isolation”), and viral shedding by detection of nucleic acids by RT-PCR (“viral shedding”).

## Methods

We conducted a rapid review of the literature using the methods outlined in the National Collaborating Centre for Methods and Tools Guidebook [8]. We used the PRISMA guidelines to report our rapid review [9]. We did not publish or pre-register a protocol for our review. From the available data communicability duration (maximums, medians or means values) was retrieved or calculated.

### Search strategy

We searched peer-reviewed and pre-print health sciences literature databases (Ovid MEDLINE, Embase, Google Scholar, medRxiv and arXiv) and the grey literature for reports or guidelines on discontinuation of isolation for SARS-CoV-2 from international and national public health organizations (World Health Organization, European Centre for Disease Prevention and Control, US Centre for Disease Control websites). Our review included two search strategies: 1) terms for ‘SARS-CoV-2’ and ‘viral clearance/shedding,’ on May 23, 2020; and 2) terms for ‘SARS-CoV-2’ and ‘viral isolation/culture,’ on July 1, 2020. We updated the second search to July 1, 2020 because there was a paucity of studies with data from viral isolation/cultures in the previous iteration of the search and identifying the duration of viable virus through studies on viral isolation was identified as key to understanding the potential infectious period of SARS-CoV-2. All databases were searched from inception and searches were limited to English. Detailed information on search strategies undertaken in each database can be found in Supplementary Materials. The references of select high-impact articles, reports from reputable sources, and existing reviews on the topic were reviewed and missing studies were added to search results. Any missing studies identified through other sources or contacts were manually added.

### Inclusion criteria

Studies presenting primary empirical data on duration of possible communicability of SARS-CoV-2 in human populations and reported in English were included. Articles that did not report data on duration of potential communicability, either in the text, figures, or tables were excluded. Studies reporting solely on pre-symptomatic durations, incubation periods, serial intervals, or on results based on statistical modelling were excluded. Studies that used data from other investigations were excluded; any reviews were identified and their references were reviewed to add relevant primary studies. When duplicate study reports were identified (i.e. a pre-print and a peer-reviewed journal article), the most recent version was included.

### Screening process

Titles and abstracts were screened for relevance independently and in duplicate by two reviewers. Full-text review was conducted by two reviewers independently and in duplicate for articles where relevance was not readily determined from title and abstracts, and any conflicts were resolved through consensus by the two reviewers.

### Data extraction process

A draft data extraction form was developed and trialled across multiple reviewers to develop the final version. Extracted data fields included study characteristics (first author, publication status, study type, sample size), study population characteristics (age, hospitalization, disease severity), method of determining infectious period (contact transmission, viral shedding, viral isolation), type of specimen(s) collected (respiratory, other), reported durations (minimum, mean, median, maximum), whether cases without symptoms (either asymptomatic or pre-symptomatic) were reported, whether the study focused solely on duration of communicability during the convalescent phase, how measurement of duration start and end was defined, and study quality. All extracted data was reviewed by a second reviewer.

### Definitions

Sample size was defined as the total number of participants for which data on communicability period was assessed. Disease severity associated with SARS-CoV-2 infection was classified according to the following definitions: mild referred to study populations reporting no symptoms or non-serious symptoms that did not require healthcare intervention, moderate severity included participants that required acute care and/or intervention, and severe disease referred to cases that required admission to the intensive care unit, critical intervention, and/or resulted in death. Studies that included participants with mild, moderate, and severe cases were classified as “mixed”; otherwise, if they included cases that were mild/moderate or moderate/severe, they were identified as belonging in the category indicating a higher level of severity. Hospitalization status was determined based on whether the study described if participants had been hospitalized during the study period. Notably, in some jurisdictions, admission to hospital appeared to be part of routine isolation policies and so this description alone was not taken as an indicator of disease severity. Studies with children were identified if they included participants aged individuals 19 years or younger. Respiratory samples included those taken from the upper (including those identified as naso/oro-pharyngeal, nasal, throat, or saliva swabs) or lower respiratory tract (from sputum or bronchial lavage specimens).

### Assessment of study quality

An adaptation of the Mixed Methods Appraisal Tool was used to assess study quality [10]. Questions were concerned with the following: 1) the study had clear research questions or objectives; 2) the collected data allowed the study to address the stated research question; 3) the research question was aimed at understanding duration of communicability; 4) there was a complete follow-up period defined to measure duration of communicability; and 5) there was clarity in how measurements were made in terms of: a) clear delineation of when measurement of communicability period started; b) sample types collected & frequency of sample collection was outlined; c) how long patients were followed (until viral clearance, study end, hospital discharge, death); and d) method of assessment of communicability was clearly explained. The results of the study quality assessment are included in Table 1. Due to the emerging nature of this topic and paucity of evidence, we did not exclude studies from our synthesis or analysis based on study quality.

**Table 1.** Characteristics of included studies, overall and broken down by method of assessment of duration of communicability.

|  | All studies n=165 |  | Contact transmission n=1 |  | Viral isolation n=3 |  | Viral shedding n=144 |  | Viral isolation and shedding n=17 |  |
| --- | --- | --- | --- | --- | --- | --- | --- | --- | --- | --- |
| **Sample size** |  |  |  |  |  |  |  |  |  |  |
| Minimum |  | 1 |  | 2861 |  | 23 |  | 1 |  | 1 |
| Maximum |  | 2861 |  |  |  | 129 |  | 584 |  | 1061 |
| Mean (SD) |  | 71.98 (245.78) |  |  |  | 72.33 (53.40) |  | 50.56 (84.78) |  | 89.24 (252.91) |
| Median (IQR) |  | 16 (67.00) |  |  |  | 65 (53.00) |  | 15 (65.25) |  | 9 (75) |
| **Study design** | **n** | **%** | **n** | **%** | **n** | **%** | **n** | **%** | **n** | **%** |
| Case control | 2 | 1.21 | 0 | 0.00 | 0 | 0.00 | 2 | 1.39 | 0 | 0.00 |
| Case report | 37 | 22.42 | 0 | 0.00 | 0 | 0.00 | 31 | 21.53 | 6 | 35.29 |
| Case series | 104 | 63.03 | 1 | 100.00 | 2 | 66.67 | 92 | 68.89 | 9 | 52.94 |
| Clinical trial | 3 | 1.82 | 0 | 0.00 | 0 | 0.00 | 3 | 2.08 | 0 | 0.00 |
| Cohort | 15 | 9.09 | 0 | 0.00 | 0 | 0.00 | 15 | 10.42 | 0 | 0.00 |
| Cross-sectional | 4 | 2.42 | 0 | 0.00 | 1 | 33.33 | 1 | 0.69 | 2 | 11.76 |
| **Measurement start** | **n** | **%** | **n** | **%** | **n** | **%** | **n** | **%** | **n** | **%** |
| Hospital admission | 37 | 22.42 | 0 | 0.00 | 0 | 0.00 | 34 | 23.61 | 3 | 17.65 |
| Date of 1<sup>st</sup> positive test | 11 | 6.67 | 0 | 0.00 | 0 | 0.00 | 11 | 7.64 | 0 | 0.00 |
| Symptom onset | 89 | 53.94 | 0 | 0.00 | 3 | 100.00 | 74 | 51.39 | 12 | 70.59 |
| Other | 28 | 16.97 | 1 | 100.00 | 0 | 0.00 | 25 | 17.36 | 2 | 11.76 |
| **Measurement end** | **n** | **%** | **n** | **%** | **n** | **%** | **n** | **%** | **n** | **%** |
| Negative test | 114 | 69.09 | 0 | 0.00 | 0 | 0.00 | 108 | 75.00 | 6 | 35.29 |
| Discharge/death | 9 | 5.45 | 0 | 0.00 | 0 | 0.00 | 7 | 4.86 | 2 | 11.76 |
| Last positive test | 6 | 3.64 | 0 | 0.00 | 0 | 0.00 | 5 | 3.47 | 1 | 5.88 |
| Other | 36 | 21.82 | 1 | 100.00 | 3 | 100.00 | 24 | 16.67 | 8 | 47.06 |
| **Continent** | **n** | **%** | **n** | **%** | **n** | **%** | **n** | **%** | **n** | **%** |
| Asia | 135 | 81.81 | 1 | 100.00 | 0 | 0.00 | 127 | 88.19 | 7 | 41.18 |
| Australia | 1 | 0.61 | 0 | 0.00 | 0 | 0.00 | 1 | 0.69 | 0 | 0.00 |
| Europe | 17 | 10.3 | 0 | 0.00 | 3 | 100.00 | 8 | 5.56 | 6 | 35.29 |
| Middle East | 2 | 1.21 | 0 | 0.00 | 0 | 0.00 | 2 | 1.39 | 0 | 0.00 |
| North America | 10 | 6.06 | 0 | 0.00 | 0 | 0.00 | 6 | 4.17 | 4 | 23.53 |
| **Age group** | **n** | **%** | **n** | **%** | **n** | **%** | **n** | **%** | **n** | **%** |
| Children | 18 | 10.91 | 0 | 0.00 | 1 | 33.33 | 17 | 11.81 | 0 | 0.00 |
| Adults | 118 | 71.52 | 0 | 0.00 | 2 | 66.67 | 102 | 70.83 | 14 | 82.35 |
| Mixed | 29 | 17.58 | 1 | 100.00 | 0 | 0.00 | 25 | 17.36 | 3 | 17.65 |
| **Hospitalization status** | **n** | **%** | **n** | **%** | **n** | **%** | **n** | **%** | **n** | **%** |
| Hospitalized | 140 | 84.85 | 0 | 0.00 | 3 | 100.00 | 124 | 86.11 | 14 | 82.35 |
| Non-hospitalized | 4 | 2.42 | 0 | 0.00 | 0 | 0.00 | 4 | 2.78 | 0 | 0.00 |
| Mixed | 15 | 9.09 | 0 | 0.00 | 0 | 0.00 | 12 | 8.33 | 2 | 11.76 |
| Unclear | 6 | 3.63 | 1 | 100.00 | 0 | 0.00 | 4 | 2.78 | 1 | 5.88 |
| **Disease severity** | **n** | **%** | **n** | **%** | **n** | **%** | **n** | **%** | **n** | **%** |
| Mild | 38 | 23.03 | 0 | 0.00 | 0 | 0.00 | 35 | 24.31 | 3 |  |
| Moderate | 35 | 21.21 | 0 | 0.00 | 0 | 0.00 | 31 | 21.53 | 4 |  |
| Severe | 13 | 7.88 | 0 | 0.00 | 1 | 33.33 | 10 | 6.94 | 2 |  |
| Mixed | 47 | 28.48 | 0 | 0.00 | 1 | 33.33 | 41 | 28.47 | 5 |  |
| Unclear | 32 | 19.39 | 1 | 100.00 | 1 | 33.33 | 27 | 18.75 | 3 |  |
| **Sample type** | **n** | **%** | **n** | **%** | **n** | **%** | **n** | **%** | **n** | **%** |
| Non-respiratory only | 3 | 1.82 | 0 | 0.00 | 0 | 0.00 | 2 | 1.39 | 1 | 5.88 |
| Respiratory + other | 161 | 97.57 | 0 | 0.00 | 3 | 100.00 | 142 | 98.61 | 16 | 94.12 |
| No sample | 1 | 0.61 | 1 | 100.00 | 0 | 0.00 | 0 | 0.00 | 0 | 0.00 |
| **Focused on Convalescent Period Only** | **n** | **%** | **n** | **%** | **n** | **%** | **n** | **%** | **n** | **%** |
| Yes | 7 | 4.24 | 0 | 0.00 | 0 | 0.00 | 6 | 4.17 | 1 | 5.88 |
| No | 158 | 95.76 | 1 | 100.00 | 3 | 100.00 | 138 | 95.83 | 16 | 94.12 |
| **Includes Asymptomatic or Pre-Symptomatic cases** | **n** | **%** | **n** | **%** | **n** | **%** | **n** | **%** | **n** | **%** |
| Yes | 73 | 44.24 | 1 | 100.00 | 0 | 0.00 | 68 | 47.22 | 4 | 23.53 |
| No | 92 | 55.76 | 0 | 0.00 | 3 | 100.00 | 76 | 52.78 | 13 | 76.47 |
| **Publication status** | **n** | **%** | **n** | **%** | **n** | **%** | **n** | **%** | **n** | **%** |
| Peer-reviewed journal | 139 | 84.24 | 1 | 100.00 | 1 | 33.33 | 121 | 84.03 | 16 | 94.12 |
| Pre-print database | 26 | 15.76 | 0 | 0.00 | 2 | 66.67 | 23 | 15.97 | 1 | 5.88 |
| **Study Quality** | **n** | **%** | **n** | **%** | **n** | **%** | **n** | **%** | **n** | **%** |
| 0 quality concerns | 30 | 18.18 | 1 | 100.00 | 1 | 33.33 | 26 | 18.06 | 2 | 11.76 |
| 1 quality concern | 43 | 26.06 | 0 | 0.00 | 1 | 33.33 | 37 | 25.69 | 5 | 29.41 |
| 2 quality concerns | 52 | 31.51 | 0 | 0.00 | 1 | 33.33 | 45 | 31.25 | 6 | 35.29 |
| 3 quality concerns | 32 | 19.39 | 0 | 0.00 | 0 | 0.00 | 28 | 19.44 | 4 | 23.53 |
| 4 quality concerns | 6 | 3.63 | 0 | 0.00 | 0 | 0.00 | 6 | 4.17 | 0 | 0.00 |
| 5 quality concerns | 2 | 1.21 | 0 | 0.00 | 0 | 0.00 | 2 | 1.39 | 0 | 0.00 |
| IQR, interquartile range; SD, standard deviation |  |  |  |  |  |  |  |  |  |  |

### Analysis

All data processing and analysis was conducted using the statistical programming language R (version 4.0.0) [11]. For studies where more than one value was reported for duration (ie, when there was data reported for multiple sample types, or results were presented in a stratified manner), the values corresponding to the higher duration of communicability were included for analysis. As analyses were generally aimed at identifying the maximum reported period of communicability, these values were pulled from each study and summarized. Raw data are available in Supplementary Materials.

## Results

### Results of literature search

From database manual searches, 2,174 records were retrieved, and 60 additional studies were identified from reference chaining and manually (2,234 total), of which 1,458 remained after removing duplicate records. Of these, 1,234 were excluded in screening and 59 in full-text review as they did not report data on duration of infectiousness, used secondary data from the literature, full-text was not accessible, or they were duplicate reports of the same study. 165 studies were included in the final synthesis. See Figure 1 for the PRISMA flowchart diagram that illustrates the study selection process.

**Figure 1.**
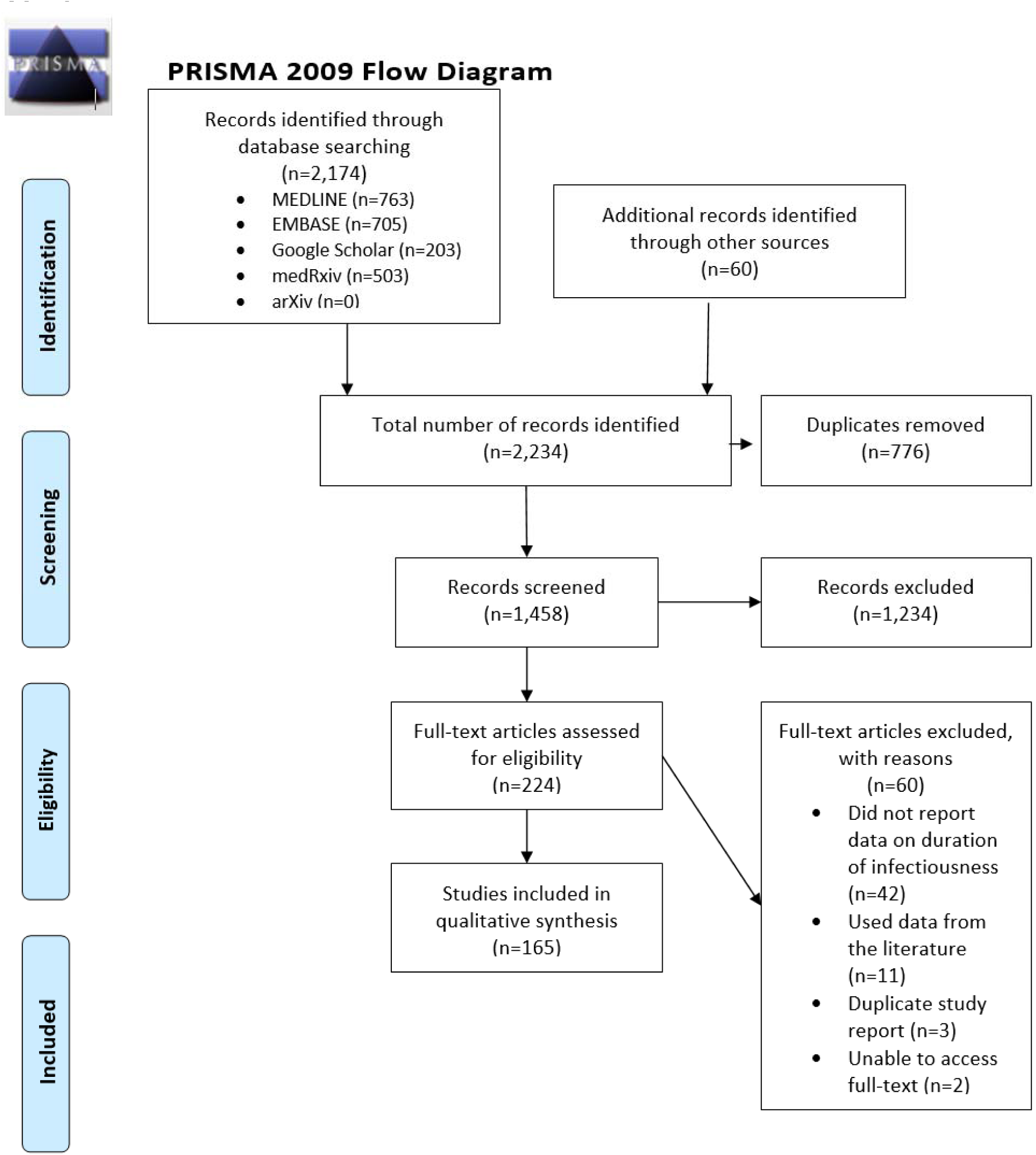
PRISMA flowchart diagram of included studies. The PRISMA flowchart details the number of studies identified through our search, and then included for analysis after screening for eligibility.

### Study characteristics

Information on study characteristics is presented in Table 1, overall and broken down by method of assessment. Out of 165 total included studies, one study investigated contact transmission, three investigated viral isolation, 144 investigated viral shedding, and 17 studies focused on both viral shedding and viral isolation. The study sample sizes of included studies ranged from 1 to 2861, with a mean of 71.98 (SD 245.78) and a median of 16 (IQR 67.00). Studies that investigated both viral isolation and shedding had a mean sample size of 89.24 (SD 252.91) and a median of 9 (IQR 65.25) (range 1-1061). The mean and median sample size in viral isolation studies was 72.33 and 65 respectively (SD 53.40; IQR, 53.00; range 23-129). In viral shedding studies, the mean sample size was 50.56 (SD 85.78) and the median was 15 (IQR 65.25) (range 1-584). The majority of studies consisted of case series (n=104, 63.03%), while 37 (22.42%) were case reports, cohort (n=15, 9.09%), cross-sectional (n=4, 2.42%), clinical trial (n=3, 1.82%) and case control studies (n=2, 1.21%) (Table 1).

The majority of studies were conducted in Asia (n=135, 81.81%), with 17 studies conducted in Europe (10.30%), 10 in North America (6.06%), 2 in the Middle East (1.21%), and 1 in Australia (0.61%) (Table 1).

Unique countries that were represented were: Australia (n=1), Canada (n=3), China (n=106), Finland (n=1), France (n=5), Germany (n=3), Hong Kong (n=4), India (n=1), Italy (n=4), Japan (n=2), Lebanon (n=1), Saudi Arabia (n=2), Scotland (n=1), Singapore (n=4), South Korea (n=7), Spain (n=1), Taiwan (n=4), Thailand (n=1), The Netherlands (n=1), United Arab Emirates (n=1), United States (n=7), and Vietnam (n=4).

Study populations had mixed disease severity (i.e., included mild, moderate and severe disease) in 47 studies (28.48%). Thirty-eight studies (23.03%) had participants with only mild symptoms, while 35 (21.21%) studies had moderate symptoms, and 13 (7.88%) with severe disease. Thirty-two studies (19.39%) did not provide information on disease severity. The majority of studies included only hospitalized patients (n=140, 84.85%). Fifteen studies (9.09%) included a mixed population of hospitalized and non-hospitalized patients, four included only non-hospitalized patients (2.42%) and for six studies (3.63%), hospitalization status of study participants was unclear. Most of the studies included only adults (n=118, 71.52%), while 18 studies (10.91%) focused solely on pediatric populations, and 29 studies (17.58%) had a mixed population of children and adults.

The starting points for measuring duration were symptom onset (n=89; 53.94%), hospital admission (n=37; 22.42%), when the patient first tested positive for SARS-CoV-2 (n=11; 6.67%), or other (n=28, 16.97%). Overall across studies, the end points of duration were the date of a single or consecutive negative RT-PCR test (n=114; 69.09%), discharge/death (n=9; 5.45%) last positive test (n=6; 3.64%), or other (n=36, 21.82%). The end point for measuring duration of viral isolation was the last reported day that virus was able to be successfully isolated and cultured (captured under “other”). For the purposes of this review, we will refer to the start of measurement of duration as ‘symptom onset’ and measurement end as ‘viral clearance.’

The majority of included studies collected data from respiratory samples, with or without another type of sample (n=161, 97.57%). Three studies (1.82%) investigated non-respiratory samples only, and 1 study (0.61%) collected no samples as it used contact transmission to determine duration of communicability.

Seventy-three studies (44.24%) included asymptomatic or pre-symptomatic positive cases. Of the studies assessing viral isolation, four studies (25.00%) also isolated and cultured virus from patients who were asymptomatic, or during their pre-symptomatic or convalescent disease period [12–15]. Two studies with data on viral isolation focused on isolating virus from non-respiratory samples, one in urine that was isolated on day 12 [16] and one in fecal matter that was isolated on day 19 after symptom onset [17]. Seven studies (4.24%) focused solely on convalescent patients who re-tested positive via RT-PCR after discharge from hospital/clearance of isolation, but only 1 of these studies attempted viral isolation, which was unsuccessful [18].

At the time of writing, 26 (15.76%) of the studies included in this review are pre-prints and thus have not undergone peer-review. Many of the remaining studies published in peer-review journals are letters to the editor or other short communications that do not fully report their methods. There was variation in study quality in both pre-prints and studies published in peer-reviewed journals. Forty-three studies (26.06%) had one study quality concern, 52 studies (31.51%) had two study quality concerns, 32 studies (19.39%) had three study quality concerns, six studies (3.63%) had four study quality concerns and two studies (1.21%) had all five study quality concerns. Only 30 studies (18.18%) did not have a single quality concern based on our assessed criteria.

Further analysis was restricted to the 155 studies with data collected from respiratory samples and that focused on the overall period of communicability, rather than solely on the convalescent period.

### Maximum reported durations of communicability

Out of 13 studies that successfully isolated and cultured virus from respiratory samples, the longest period that viable virus was able to be isolated from a case was 32 days after symptom onset; the median and mean durations for longest period of viral isolation across studies were 9 days (IQR 10) and 11.8 days (SD 8.47), respectively (Table 2). From the 133 studies with data on maximum duration of viral shedding from respiratory samples, the shortest reported time until viral clearance based on RT-PCR test results was five days from symptom onset, while the maximum was 95 days, with respective median and mean values of 24 and 28.7 days (IQR 19; SD 15.80). One study presented data on infectious period based on contact transmission; this study found that the longest time since symptom onset in the index case from which a secondary case was infected was five days [19].

**Table 2.** Summary statistics on duration of maximum duration of communicability from studies reporting on contact transmission, isolation and culture of live virus from respiratory samples, and detection of viral shedding by RT-PCR from respiratory samples. Note that some studies provided data for both viral isolation and viral shedding.

| Method of analysis | Number of included studies | Shortest maximum reported duration | Mean maximum reported duration (SD) | Median maximum reported duration (IQR) | Longest maximum reported duration |
| --- | --- | --- | --- | --- | --- |
| Contact Transmission | 1 | n/a | n/a | n/a | 5 |
| Viral Isolation | 13 | 2 | 11.8 (8.47) | 9 (10) | 32 |
| Viral Shedding | 133 | 5 | 28.7 (15.80) | 24 (19) | 95 |
| IQR = interquartile range; SD = standard deviation |  |  |  |  |  |

### Findings from viral isolation studies

To assess how long viable virus could be obtained, considered as a proxy measure of infectious potential, we focused specifically on studies that investigated isolation and culturing of virus from respiratory samples (n=16). Studies represented a mix of study designs (case reports, case series, and cross-sectional studies) and a wide range of sample sizes from which viral isolation was attempted (range 1-915) and/or successful (range 1-204). Studies determined duration of viral viability either by taking a cross-section of diagnostic samples collected at different times from symptom onset or by serially collecting samples from the same individuals over time; not all studies attempted viral isolation until viral clearance. Three studies were unable to isolate and culture any virus (Table 3).

**Table 3.** Detailed data from papers investigating durations until viral isolation and culture. Papers are presented in alphabetical order by first author.

| First Author<br>Journal & publication date/status | Country | Study design | Study population | Sample Size | Sample types taken for isolation, sampling method | Viral shedding (max) | Viral isolation (max) | Notable findings | Symptoms |
| --- | --- | --- | --- | --- | --- | --- | --- | --- | --- |
| Arons et al.(12)<br>*New England Journal of Medicine* (April 24, 2020) | United States | Serial cross-sectional | Patients in a skilled nursing facility with mixed disease severity, a mean age of 78.6, 98% had a co-morbidity | 48 (27 individuals with 2 samples taken) | NP & OP samples, collected at 2 time points, 1 week apart. | 13 days | 9 days | Positive cultures in 17 out of 24 pre-symptomatic patents and 1 out of 3 asymptomatic patients.<br>Viable virus isolated up to 6 days before onset of symptoms. RT-PCR positive up to 7 days prior to symptom onset.<br>RT-PCR Ct values ranged from 13.7-37.9 in positive samples. | Includes asymptomatic and pre-symptomatic patients. Most common new symptoms were fever, cough and malaise. |
| Bullard et al. (26)<br>*Clinical Infectious Diseases* (May 22, 2020) | Canada | Cross-sectional | All samples in this study were obtained to support routine care and surveillance of the public health response in the province of Manitoba, Canada. All suspect COVID-19 cases had SARS-CoV-2 RT-PCR performed on nasopharyngeal (NP) or endotracheal (ETT) samples at Cadham Provincial Laboratory. | 90 (26 with positive viral isolation ) | NP and endotracheal samples, from diagnostic samples of individuals who tested positive by RT-PCR from day 0 to 21 post symptom onset | 21 days | 8 days | Positive viral culture samples had significantly lower Ct values than negative cultures (17 (16-19) vs 27 (22-33)). For every increase in unit in Ct value, the odds of a positive culture decreased by 32%.<br>Time from symptom to test time was significantly lower in positive vs negative cultures (3 (2-4) vs 7 (2-11)). For every day | Not described. |
|  |  |  | Median age of the patients sampled was 45 (range: 30-59 years). Forty nine percent of samples were from males. |  |  |  |  | since symptom the odds of a positive culture decreased by 37%.<br>Probability of a successful positive culture peaked on day 3 and decreased after that point.<br>No growth in samples with Ct >24. |  |
| Chang et al. (42)<br>*The Journal of Allergy and Clinical Immunology in Practice* (June 20, 2020) | China | Case series | Hospitalized patients with mixed disease severity, who re-tested positive after discharge. 3 required ICU admission. | 69 (4 attempted and 0 positive viral isolations) | Throat samples, collected serially while in hospital and after discharge. Only attempted viral isolation from samples taken during the convalescent period, sent while patients were at home. | 57 days | N/A | No positive viral isolation/culture during convalescent period. | Fever present in 82% of patients, cough in 60% of patients, sputum in 25%. 2 patients were asymptomatic. |
| Decker et al. (15)<br>*American Journal of Transplantation* (June, | Germany | Case report | 62 year old male heart transplant recipient who was hospitalized with mild disease severity | 1 | Throat samples, collected serially at 10 time points until day 35 of illness. | >35 days (patient still testing positive at study end) | 21 days | Patient post-symptomatic at time of positive viral cultures. | Fever on day 1 that resolved within 12 hours, second fever that spiked and resolved at day 7. Other symptoms were mild rhinorrhea and |
| 2020) |  |  |  |  |  |  |  |  | impaired exercise capacity. |
| Folgueira et al. (28)<br>*MedRxiv\* Preprint (June 12, 2020)* | Spain | Cross-sectional | Patients with mixed disease severity. Patients with mild symptoms (n=24) were healthcare workers, largely women, and assessed in outpatient settings. Patients with severe disease were hospitalized (n=41), of which 5 died. | 65 (106 samples taken) | NP samples and bronchial aspirates, from diagnostic samples of individuals diagnosed as outpatients and those followed up for hospital care. Median time of sample collection for hospitalized patients was 19.6 days and for outpatients was 16.5 days. | 32 days | N/A | Positive viral culture was obtained from a higher proportion of patients with severe disease vs mild symptoms (53.4% of samples vs 36.0%). All samples from deceased patients were able to be cultured.<br>Samples with Ct values < 25 had >90% positive viral culture. However, samples with low viral loads (Ct > 35) could still harbor viable virus. | 6 hospitalized patients were admitted to the ICU and required mechanical ventilation. 5 patients died. |
| Kumar et al. (13)<br>*Canadian Medical Association Journal (April 24, 2020)* | Canada | Case report | 76 year old man with multiple comorbidities, who initially tested negative for COVID-19. | 1 | NP sample, collected on day 4 of hospital admission (which was 11 days after exposure, and day 6 after cough worsened) | 17 days | N/A | No positive viral isolation/culture.<br>Sputum culture conducted on day 4, which is the same day that patient tested negative for SARS-CoV-2 in a NP swab. NP swab, positive on day 7 of admission (18 days after exposure; 13 days after cough worsened) | Worsening cough on days 1-6, fatigue, exertional dyspnea, fevers, low appetite and diarrhea on days 2-6 |
| Gautret et al. (43)<br>*Travel Medicine and Infectious diseases (April 2020)* | France | Case series | Hospitalized patients with age range of 18 to 88 years, 57.5% had at least one comorbidity. 3 patients were transferred to ICU, 1 patients died. | 80 (53 with positive viral isolation ) | NP samples, collected daily beginning at treatment. | 12 days | 9 days | Viral cultures were negative in 97.5% of patients at day 5. | 4 patients were asymptomatic, 14 patients had fever as a symptom, 33 had upper respiratory tract symptoms and 43 had lower respiratory tract symptoms. |
| Haveri et al. (24)<br>*Euro Surveillance (Mar 25, 2020)* | Finland | Case report | First COVID-19 case in Finland. Hospitalized woman in her 30s from Wuhan with mild disease severity. | 1 | NP samples, collected serially, on days 3, 4, 9, 10, 20 and 23, unclear when viral isolation was attempted. | 8 days | 4 days | Showed late seroconversion: Antibodies were undetectable on Day 4 after onset of symptoms, IgG titres rose to 80 and 1,280 and IgM titres to 80 and 320 on Days 9 and 20.<br>Ct values on day 4 for different RT-PCR targets: E (29.59), RdRp (30.87), N (31.78). | Respiratory symptoms on onset, followed by high fever on day 2, post-symptomatic by day 7. |
| Kim et al. (25)<br>*J Korean Med Sci (Feb 24, 2020)* | South Korea | Case series | First 2 patients with COVID-19 in South Korea. 1 35-year old woman and 1 55-year old man, both with mild symptoms. | 2 | Respiratory samples, collected daily starting day 2 and day 14 of illness. | 26 days | N/A | No positive viral isolation/ culture.<br>Upper respiratory RdRp Ct values ranged from 25.05-36.69 and lower respiratory RdRp Ct values ranged from 22.05-32.63. | Patient 1 had fever at onset of illness until day 9, nasal congestion on days 6-8, dyspnea on days 9-11, cough on days 7-13, sputum on days 9-13 and mild loose stools on days 4-19. Patient 2 had sore throat from illness onset until day 17, fever beginning around day 10 and lasting until 16, cough on days 16-18 and loose stool on days 19-21. |
| Kujawski et al. (27)<br>*Nature (June 26, 2020)* | United States | Case series | Convenience sample of the first 12 US patients confirmed to have COVID-19. 5 patients had underlying conditions. Median age was 53 years (range: 21–68); 8 patients were male. Had mild to moderate illness with 7 patients hospitalized but none requiring mechanical ventilation and all showing recovery. | 12 (9 with positive viral isolation ) | NP and OP samples, which were taken on days 1-9 from symptom onset. Not attempted in later specimens. | 29 days | 9 days | Positive viral isolation from samples with RT-PCR Ct values of 12.3–35.7. | Median symptom duration of 14 days, cough reported as last symptom to resolve. Median duration of fever was 9 days (range 2-11), with a peak body temperature at median 9 days (range 4-10) |
| Lescure et al. (23)<br>*Lancet Infectious Diseases (Mar 27, 2020)* | France | Case series | Patients were three men (aged 31 years, 48 years, and 80 years) and two women (aged 30 years and 46 years). | 5 | NP samples, taken from patients once only at days 2, 2, 6, 7, 9 since symptom onset. | 24 days (until patient death) | 2 days | Positive viral isolation in samples with RdRp Ct values of 23.6 and 24.4, E gene Ct of 22.8 and 20.0, RdRp IP Ct of 23.0 and 19.3, GAPDH (housekeeping gene) Ct of 26.5 and 25.6.<br>Positive isolate titre was $6.25 \times 10^5$ and $3.0 \times 10^7$. | Describes 3 subtypes of symptom progression: first, mild cases through two paucisymptomatic patients aged younger than 50 years who were diagnosed early, with high viral load in nasopharyngeal samples, suggesting a significant shedding of SARS-CoV-2, reflected by virus detection by RT-PCR; second, two young patients presenting with mild symptoms at admission and experiencing a secondary progression to pneumonia and severe disease by days 10–11; and third, an<br>older patient with a rapid evolution towards critical disease with multiple organ failure and a long and sustained persistence of SARS-CoV-2 nasopharyngeal detection associated with viral RNA detection in multiple sites |
| L'Huillier et al (22)<br>*Emerging Infectious Diseases (June 30, 2020)* | Switzerland | Case series | Children with mild disease. Isolated virus from children of all ages; the youngest was 7 days of age | 23 (12 with positive viral isolation ) | NP samples, taken from patients at days 0 - 5 after symptom onset. Did not attempt after day 5. | N/A | 5 days | Median viral load was higher for patients with positive viral isolation than for those without isolation. ($1.7 \times 10^8$ vs $6.9 \times 10^3$). | Symptoms did not differ between children from whom a positive viral isolation was obtained vs not. |
| Liu et al. (20)<br>*The Journal of Infection (April 18, 2020)* | Taiwan | Case report | 50 year old hospitalized woman with mild disease and no comorbidities | 1 | Throat and sputum samples, collected daily. | 63 days | 18 days | Viral cultures positive after resolution of symptoms. Antibodies detected on day 10, but viral cultures remained positive until day 18.<br>Virus isolated from throat swabs at admission, and from sputum until 18 days after symptom onset. | Fever resolved on day 10. |
| Million et al. (14)<br>*Travel* | France | Case series | Hospitalized patients with a mean age of 47.9 (SD 17.5), 2.6% had cancer, 7.4% had | 1061 (915 attempted, 204 | NP samples, collected daily for 11 participants. | >15 days (patient still testing | 9 days | Prolonged viral shedding associated with high viral load at diagnosis, but no positive viral cultures after | Not described. |
| *Medicine & Infectious Disease (May, 2020)* |  |  | diabetes, 4.3% had coronary artery disease, 14% had hypertension, 10.5% had chronic respiratory disease and 5.8% had obesity. 973 patients (91.7% had good clinical outcome) with 38 severe outcomes including death. | positive viral isolations, 11 individuals with daily samples) |  | positive at study end) |  | day 9. |  |
| van Kampen et al. (21)<br>*MedRxiv* *Pre-print (June 9, 2020) | The Netherlands | Case series | Hospitalized patients with severe or critical COVID-19, admitted to medium acute care (40, 31%) or to intensive care (89, 69%). 11 patients were severely immunocompromised and 19 were non-severely immunocompromised. | 129 (23 individuals with positive viral isolations) | Upper respiratory and sputum samples, collected serially from diagnostic samples only, not at pre-defined intervals. | N/A | 20 days | Median duration of infectious virus shedding was 8 days (IQR 5 – 11). ≤5% probability of isolating infectious virus when duration of symptoms was 15.3 days or more (95% CI 13.2-17.2)<br><5% probability of isolation when viral load went below 6.51 log<sub>10</sub> RNA copies per mL.<br>Viral load was associated with infectious viral shedding; median viral load was significantly higher in culture positive samples than in culture negative samples.<br>Shedding of infectious virus dropped rapidly to undetectable levels upon seroconversion. | Assumption that all patients with positive viral isolation were symptomatic at the time of sampling, given the patient population. |
| Wölfel et al.<br>(29)<br>*Nature (April 1, 2020)* | Germany | Case series | Hospitalized, young-to-middle aged patients with minimal pre-existing disease (one each of hypothyroidism, COPD, hypercholesteremia), and relatively mild symptoms. Patients identified based on close contact with an index case and not based on symptoms. | 9 | OP, NP, and sputum samples, collected daily beginning from 2-8 days from symptom onset. Isolation attempted on multiple occasions from positive samples. | 28 days | 8 days | Peak viral shedding in upper respiratory tract during first 5 days, with prolonged shedding in sputum. Sputum samples remained RNA-positive over three weeks in six of the nine patients, despite full resolution of symptoms.<br>Successful viral isolation found before and after seroconversion.<br>Threshold of at least 100,000 viral RNA copies per ml of sputum required for isolation.<br>Positive rate of virus isolation in first week of symptoms was 16.66% in NP/OP swab samples and 83.33% in sputum samples, but after day 8 no positive isolates were obtained despite ongoing high viral loads.<br>Seroconversion associated with a steady, not abrupt, decline of viral load. Seroconversion occurred for all patients by day 14. | 1 patient had fever as an initial symptom and 3 patients showed fever as a later symptom and the fever resolved in these 3 patients on day 4, day 4 and day 8. 1 patient was asymptomatic. |
| Ct, cycle threshold; E, envelope protein gene; GAPDH, glyceraldehyde-3-phosphate dehydrogenase gene (reference housekeeping gene); IgG, immunoglobulin G; IgM, immunoglobulin M; N, nucleocapsid protein gene; NP, nasopharyngeal; OP, oropharyngeal; RdRp, RNA-dependent RNA polymerase gene |  |  |  |  |  |  |  |  |  |

Four studies reported viral isolation beyond 10 days. The longest period that virus could be cultured after symptom onset was 32 days, from hospitalized patients with severe disease; this was a study, comparing duration of viral culture from patients with mild or severe disease. A duration of 21 days was identified from a report of a single individual with mild symptoms but with significant underlying conditions including a recent heart transplant [15]. A separate case report of an individual without any significant comorbidities and also with mild disease symptoms reported viable virus until 18 days [20]. One case series of 129 hospitalized patients with moderate to severe disease, and mixed degrees of comorbidities, reported a maximum duration of viable virus of 20 days, with a median time to viral clearance of eight days [21].

The relationship between viral viability and symptom presentation was inconsistent. Some studies reported successful viral isolation from individuals who did not have symptoms at the time of sample collection [12,15,20,22], others isolated viable virus from individuals who were symptomatic at the time of sample collection [23,24], while other studies were unable to isolate virus from symptomatic individuals [13,25] (Table 3). There were inconsistent data available on viral load and/or RT-PCR cycle threshold (Ct) values and duration of viability, though a number of studies reported an upper limit on these measures for virus viability [12,22,23,25–28]. Few studies investigated seroconversion status and viral viability; one reported that seroconversion was associated with an inability to isolate virus [28] while others showed that virus continued to be isolated after detection of antibodies [21, 29]. A case report found that viral culture was successful while antibodies were not detectable, with seroconversion occurring later [24]. Finally, one study focused on pediatric populations reported viral viability from children as young as seven days of age [22].

### Findings from studies investigating both viral isolation and viral shedding

A subset of these studies was further analyzed to compare the duration of viral nucleic acid shedding against the duration of virus viability. There were nine studies that provided data from respiratory samples on the duration of both viral nucleic acid shedding assessed by RT-PCR as well as viral isolation. All studies reported persistent viral shedding after virus was no longer able to be isolated and cultured (Figure 1). Across these studies, the median duration after symptom onset that virus was successfully isolated and cultured was 9 days, while the corresponding median value for longest duration until viral clearance by RT-PCR was 24 days (Figure 2).

**Figure 2.**
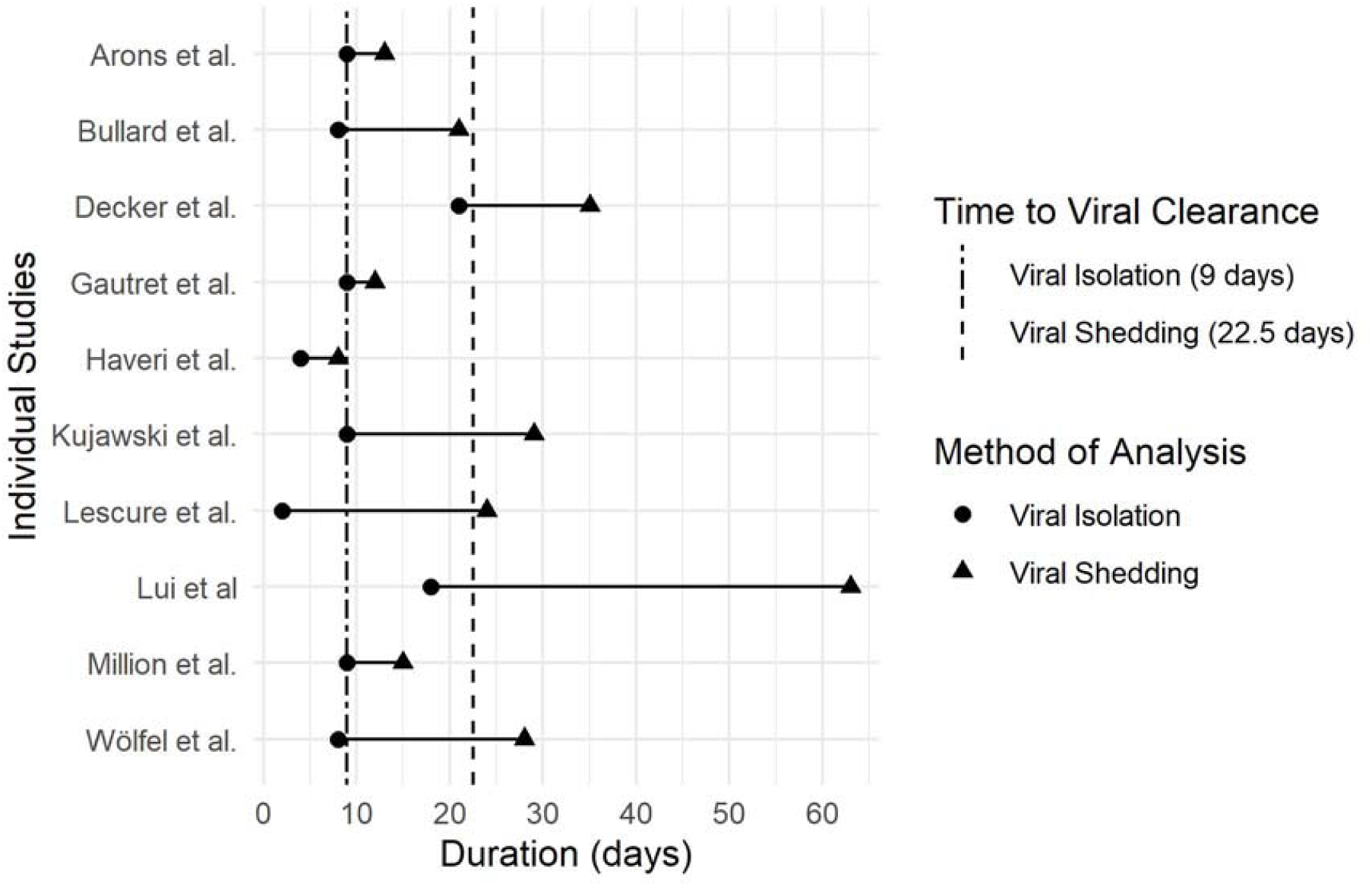
Maximum reported durations of communicability from studies (n=10) with data on viral isolation and viral shedding in respiratory SARS-CoV-2 samples. Each horizontal line represents data from the same study, with points indicating the longest time to viral clearance reported from each study for viral isolation (circles) and viral shedding (triangles). Vertical lines represent median reported values across all studies on longest time to viral clearance for viral isolation (dashed and dotted line) and viral shedding (dashed line).

### Findings from viral shedding studies with data on both maximum and median (or mean) durations of communicability

Finally, we explored how long it might take to achieve viral clearance based solely on detection of viral nucleic acid by RT-PCR tests. This analysis was specific to studies that reported measures of both central tendency (median, or mean when median was not reported) and maximum durations until viral clearance. Measures of central tendency can help to understand the population distribution in contrast to the maximum reported durations which are taken from a single data point/individual and thus more sensitive to outliers. Similarly, focusing on studies with multiple participants may provide more robust measures of time to viral clearance compared to case reports presenting data on single outlying individuals. Across these studies (n = 63), measurements of central tendency reported a median time to viral clearance of 15 days, while the median measurement for the longest reported time to viral clearance across all studies was 27 days (Figure 3).

**Figure 3.**
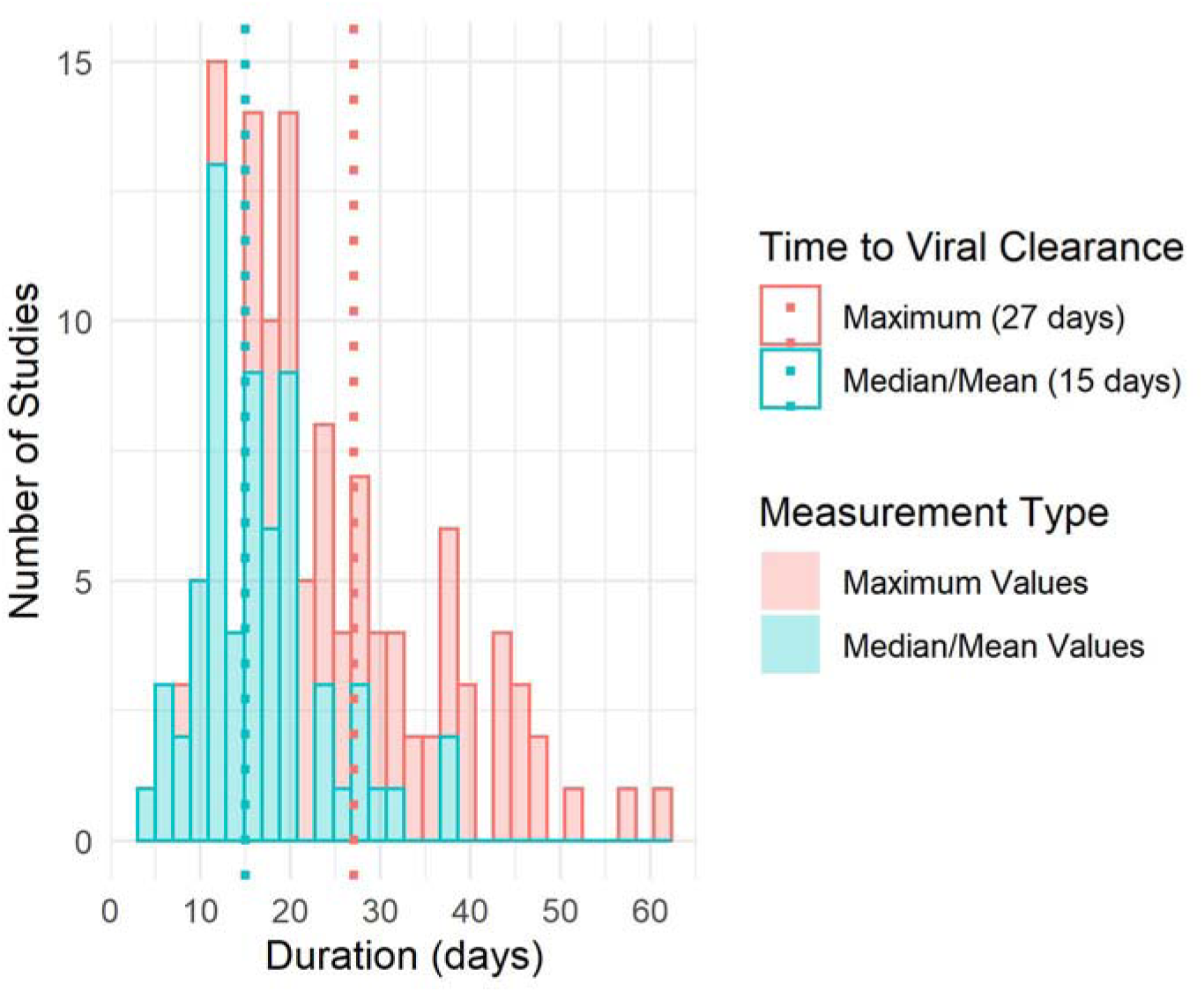
Histograms displaying the distribution of durations of communicability from studies reporting data on both median/mean and maximum durations to viral clearance (n=63). Data on median/mean values are shown in teal and maximum values are shown in pink. Dotted vertical lines refer to median values across all studies for median/mean duration (teal) and maximum duration (pink) until viral clearance.

We also stratified analyses by disease severity; however, data were limited there were only 30 studies where study populations could be identified as having had mild, moderate, or severe disease (Supplementary Figure 1). Fourteen studies each had populations with mild and moderate disease severity, while only two had populations with severe disease. Mean values for duration of median/mean viral shedding were 14 days (IQR 8.13), 15.5 days (IQR 9.25), and 24 days (IQR 7) for mild, moderate, and severe disease respectively. The median values for longest duration viral shedding from studies with mild, moderate, and severe disease were 20 (IQR 8.25), 26.5 (IQR 14), and 47.5 days (IQR 0.50).

## Discussion

The results of this review present information relevant for policies on discontinuation of isolation for individuals infected with SARS-CoV-2. However, findings should be interpreted in light of significant limitations. There was a general paucity of high-quality evidence. Several of the included studies did not follow participants until viral clearance or collect samples at consistent intervals throughout the communicable period. We identified few studies that investigated viral isolation and many of the included studies had small sample sizes, making it difficult to draw robust findings from the n-of-1 studies. The included studies suffer from selection bias and non-representative populations, as case reports and case series often focus on highly specific clinical populations and may bias toward incidences of prolonged viral shedding.

Findings from the available evidence support the general guidance of a minimum 10 day period of isolation [31]. Most studies showed an inability to isolate and culture virus beyond nine days. However, there should be additional precautions and longer observation considered for individuals being released into high-risk institutional settings such as long-term care facilities or those remaining in in-patient units, given that viable virus has been reported at 18 [30], 20 [21], 21 [15], and 32 days [28]. Notably, these positive findings were from individuals with mixed levels of underlying comorbidities, and who presented with heterogeneous disease profiles, from mild symptoms to severe complications and death associated with infection with SARS-CoV-2. An additional impetus for taking a precautionary approach is the inconsistency from studies reporting on symptomatology and viable virus; while some studies were not able to successfully culture virus from asymptomatic or post-symptomatic individuals [13,14], others successfully cultured virus from individuals who were not symptomatic at the time of specimen collection [12,15].

Findings from this review also raise some limitations of fully relying on a RT-PCR-based strategy. Exploration of the potential duration of viral clearance by RT-PCR tests yielded a median duration of 15 days until viral clearance, which is in line with guidelines suggesting a 10 – 14 day isolation period. However, studies that presented data on both viral shedding and viral isolation demonstrate that individuals may continue to test positively on nucleic acid tests, after they are no longer infectious. In exceptional cases, viral shedding may persist for several weeks [18,20,32–36].

While viral isolation and culture is a better proxy for understanding communicable period than detection of viral nucleic acid, it is not feasible to apply at scale in routine clinical practice. A number of commentaries and studies have suggested the use of additional metrics, such as disease severity [21,28], viral load [29], and presence of antibodies [21], in conjunction with a positive RT-PCR test, to help manage the issue of persistent viral shedding [37–39]. In the current study, analysis of papers with RT-PCR findings suggested a potential trend toward longer viral shedding with more severe disease; studies on viral isolation have further suggested a longer duration of viable viral shedding [21,28]. However, available data to assess this were limited and there is a scarcity of reliable evidence. Though a fulsome analysis on this topic was not conducted, findings on the relationship between seroconversion, viral load, and viral viability were inconsistent. It is anticipated that future studies will be able to provide more clarity on these issues.

There were a number of additional limitations identified in this rapid review. The included studies revealed heterogeneity in measurements of duration, including when measurement started and criteria for determining viral clearance. Some studies failed to clearly report their methods and so were unclear about when measurement started and/or ended or did not clearly report the type of sample collected (e.g., upper or lower respiratory tract). Additionally, many studies had incomplete follow-up. While studies reported a median or maximum duration of viral shedding, some only followed patients until the end of the study period rather than to an endpoint that consisted of reliable evidence of persistent viral clearance.

Our review is also limited by the need for timely appraisal of critical evidence for policy decisions and by the rapidly evolving literature. A rapid review aims to synthesize key information in a timely fashion and necessarily involves less comprehensive search methods than a systematic review. As a result of limiting the sensitivity of our search strategy and the sources searched, we may have missed some studies. Additionally, while we attempted to minimize the inclusion of repeated participants across studies by excluding studies analyzing secondary data, we did not investigate or contact authors to determine whether participants were included in multiple study populations. Also due to time constraints, we did not conduct a fulsome assessment of study quality. We found significant variations in study quality and a more detailed appraisal of study quality may reveal additional issues. Lastly, due to the rapidly growing literature on this topic, new studies have been released since the dates of our last searches [40,41].

Despite the limitations raised above, this study is strengthened by the comprehensive and updated search strategy that was employed, the clear delineation of the question of interest and corresponding specific criteria used for study inclusion, and the standardized data extraction that enabled the analyses presented herein. The raw data have been provided to assist other research and clinical teams who may find this detailed information helpful. This research highlights the critical need to update policies and practice through robust syntheses of newly emerging evidence.

## Data Availability

Raw data are available online or upon request.

https://drive.google.com/file/d/1b0at1Szp4GHZfUkUy9eD3TZMHjB3KCf_/view?usp=sharing

## Acknowledgements

We would like to thank Kelsey Furk and John Harding for their contributions to the development and design of this study.

## SUPPLEMENTARY MATERIALS

### Appendix A – Search Terms Used

#### 1. Viral Clearance/Shedding Searches

##### MEDLINE

1. ((((exp Coronavirus/ or exp Coronavirus Infections/ or (coronavirus* or corona virus* or OC43 or NL63 or 229E or HKU1 or HCoV* or ncov* or covid* or sars-cov* or sarscov* or Sars-coronavirus* or Severe Acute Respiratory Syndrome Coronavirus*).mp.) and ((20191* or 202*).dp. or 20190101:20301231.(ep).)) not (SARS or SARS-CoV or MERS or MERS-CoV or Middle East respiratory syndrome or camel* or dromedar* or equine or coronary or coronal or covidence* or covidien or influenza virus or HIV or bovine or calves or TGEV or feline or porcine or BCoV or PED or PEDV or PDCoV or FIPV or FCoV or SADS-CoV or canine or CCov or zoonotic or avian influenza or H1N1 or H5N1 or H5N6 or IBV or murine corona*).mp.) or ((((pneumonia or covid* or coronavirus* or corona virus* or ncov* or 2019-ncov or sars*).mp. or exp pneumonia/) and Wuhan.mp.) or (2019-ncov or ncov19 or ncov-19 or 2019-novel CoV or sars-cov2 or sars-cov-2 or sarscov2 or sarscov-2 or Sars-coronavirus2 or Sars-coronavirus-2 or SARS-like coronavirus* or coronavirus-19 or covid19 or covid-19 or covid 2019 or ((novel or new or nouveau) adj2 (CoV on nCoV or covid or coronavirus* or corona virus or Pandemi*2)) or ((covid or covid19 or covid-19) and pandemic*2) or (coronavirus* and pneumonia)).mp. or COVID-19.rx,px,ox. or severe acute respiratory syndrome coronavirus 2.os. or (“32240632” or “32236488” or “32268021” or “32267941” or “32169616” or “32267649” or “32267499” or “32267344” or “32248853” or “32246156” or “32243118” or “32240583” or “32237674” or “32234725” or “32173381” or “32227595” or “32185863” or “32221979” or “32213260” or “32205350” or “32202721” or “32197097” or “32196032” or “32188729” or “32176889” or “32088947” or “32277065” or “32273472” or “32273444” or “32145185” or “31917786” or “32267384” or “32265186” or “32253187” or “32265567” or “32231286” or “32105468” or “32179788” or “32152361” or “32152148” or “32140676” or “32053580” or “32029604” or “32127714” or “32047315” or “32020111” or “32267950” or “32249952” or “32172715”).ui.)) and 20191201:20301231.(dt). (8287)
2. (infectious or transmission or transmitted).ti. (130621)
3. ((viral* or virus) adj2 (shed* or clear* or load or dynamics or detect* or assessment or duration)).tw,kf. (54897)
4. ((discharge? or recovered) adj6 positive).tw,kf. (2484)
5. ((communicable or infectious) adj2 period).tw,kf. (496)
6. or/2-5 (185826)
7. 1 and 6 (423)
8. limit 7 to english language (409)

##### Embase

1. ((exp Coronavirinae/ or coronavirus*.mp.) and (wuhan or beijing or shanghai or 2019-nCoV or Covid-19 or SARS-CoV-2).mp.) or ((Coronavirus*.ti. or (novel coronavirus*.mp. and (exp China/ or china.mp.)) or ((pneumonia.mp. or exp pneumonia/) and Wuhan.mp.) or (“Covid-19” or “2019-nCoV” or “SARS-CoV-2”).mp. or exp Coronavirus Infection/) and (“2020” or “2021”).yr.) (7113)
2. (infectious or transmission or transmitted).ti. (131541)
3. ((viral* or virus) adj2 (shed* or clear* or load or dynamics or detect* or assessment or duration)).tw,kw. (76591)
4. ((discharge? or recovered) adj6 positive).tw,kw. (3547)
5. ((communicable or infectious) adj2 period).tw,kw. (572)
6. or/2-5 (208949)
7. 1 and 6 (411)
8. limit 7 to english language (388)

##### Google Scholar

- Covid AND “viral shedding”
- Covid AND “viral clearance”
- Covid AND transmission
- Covid AND asymptomatic
- Covid AND infectious period

##### medRxiv/arXiv

- Covid viral shedding
- Covid infectious period
- Covid asymptomatic
- Covid “viral clearance”

#### 2. Viral Isolation/Culture Searches

##### MEDLINE

Database: Ovid MEDLINE(R) and Epub Ahead of Print, In-Process & Other Non-Indexed Citations, Daily and Versions(R) <1946 to July 01, 2020>

Search Strategy:

----------------------------------

1. ((((exp Coronavirus/ or exp Coronavirus Infections/ or (coronavirus* or corona virus* or OC43 or NL63 or 229E or HKU1 or HCoV* or ncov* or covid* or sars-cov* or sarscov* or Sars-coronavirus* or Severe Acute Respiratory Syndrome Coronavirus*).mp.) and ((20191* or 202*).dp. or 20190101:20301231.(ep).)) not (SARS or SARS-CoV or MERS or MERS-CoV or Middle East respiratory syndrome or camel* or dromedar* or equine or coronary or coronal or covidence* or covidien or influenza virus or HIV or bovine or calves or TGEV or feline or porcine or BCoV or PED or PEDV or PDCoV or FIPV or FCoV or SADS-CoV or canine or CCov or zoonotic or avian influenza or H1N1 or H5N1 or H5N6 or IBV or murine corona*).mp.) or ((((pneumonia or covid* or coronavirus* or corona virus* or ncov* or 2019-ncov or sars*).mp. or exp pneumonia/) and Wuhan.mp.) or (2019-ncov or ncov19 or ncov-19 or 2019-novel CoV or sars-cov2 or sars-cov-2 or sarscov2 or sarscov-2 or Sars-coronavirus2 or Sars-coronavirus-2 or SARS-like coronavirus* or coronavirus-19 or covid19 or covid-19 or covid 2019 or ((novel or new or nouveau) adj2 (CoV on nCoV or covid or coronavirus* or corona virus or Pandemi*2)) or ((covid or covid19 or covid-19) and pandemic*2) or (coronavirus* and pneumonia)).mp. or COVID-19.rx,px,ox. or severe acute respiratory syndrome coronavirus 2.os. or (“32240632” or “32236488” or “32268021” or “32267941” or “32169616” or “32267649” or “32267499” or “32267344” or “32248853” or “32246156” or “32243118” or “32240583” or “32237674” or “32234725” or “32173381” or “32227595” or “32185863” or “32221979” or “32213260” or “32205350” or “32202721” or “32197097” or “32196032” or “32188729” or “32176889” or “32088947” or “32277065” or “32273472” or “32273444” or “32145185” or “31917786” or “32267384” or “32265186” or “32253187” or “32265567” or “32231286” or “32105468” or “32179788” or “32152361” or “32152148” or “32140676” or “32053580” or “32029604” or “32127714” or “32047315” or “32020111” or “32267950” or “32249952” or “32172715”).ui.)) and 20191201:20301231.(dt). (29800)
2. ((viral* or virus) adj6 (culture? or isolation or isolated)).tw,kf. (42320)
3. 1 and 2 (103)

##### Embase

Database: Embase <1974 to 2020 July 01>

Search Strategy:

---------------------------

1. ((exp Coronavirinae/ or coronavirus*.mp.) and (wuhan or beijing or shanghai or 2019-nCoV or Covid-19 or SARS-CoV-2).mp.) or ((Coronavirus*.ti. or (novel coronavirus*.mp. and (exp China/ or china.mp.)) or ((pneumonia.mp. or exp pneumonia/) and Wuhan.mp.) or (“Covid-19” or “2019-nCoV” or “SARS-CoV-2”).mp. or exp Coronavirus Infection/) and (“2020” or “2021”).yr.) (26353)
2. ((viral* or virus) adj6 (culture? or isolation or isolated)).tw,kw. (41653)
3. 1 and 2 (106)

##### Google Scholar

- Covid virus isolation
- Covid viral isolation
- Covid virus culture
- Covid viral culture

##### medRxiv/arXiv

- Title “COVID” (match all words) and abstract or title “viral isolation” (match phrase words)
- Title “COVID” (match all words) and abstract or title “virus isolation” (match phrase words)
- Title “COVID” (match all words) and abstract or title “virus culture” (match phrase words)
- Title “COVID” (match all words) and abstract or title “viral culture” (match phrase words)

### Appendix B – Inclusion/Exclusion Criteria

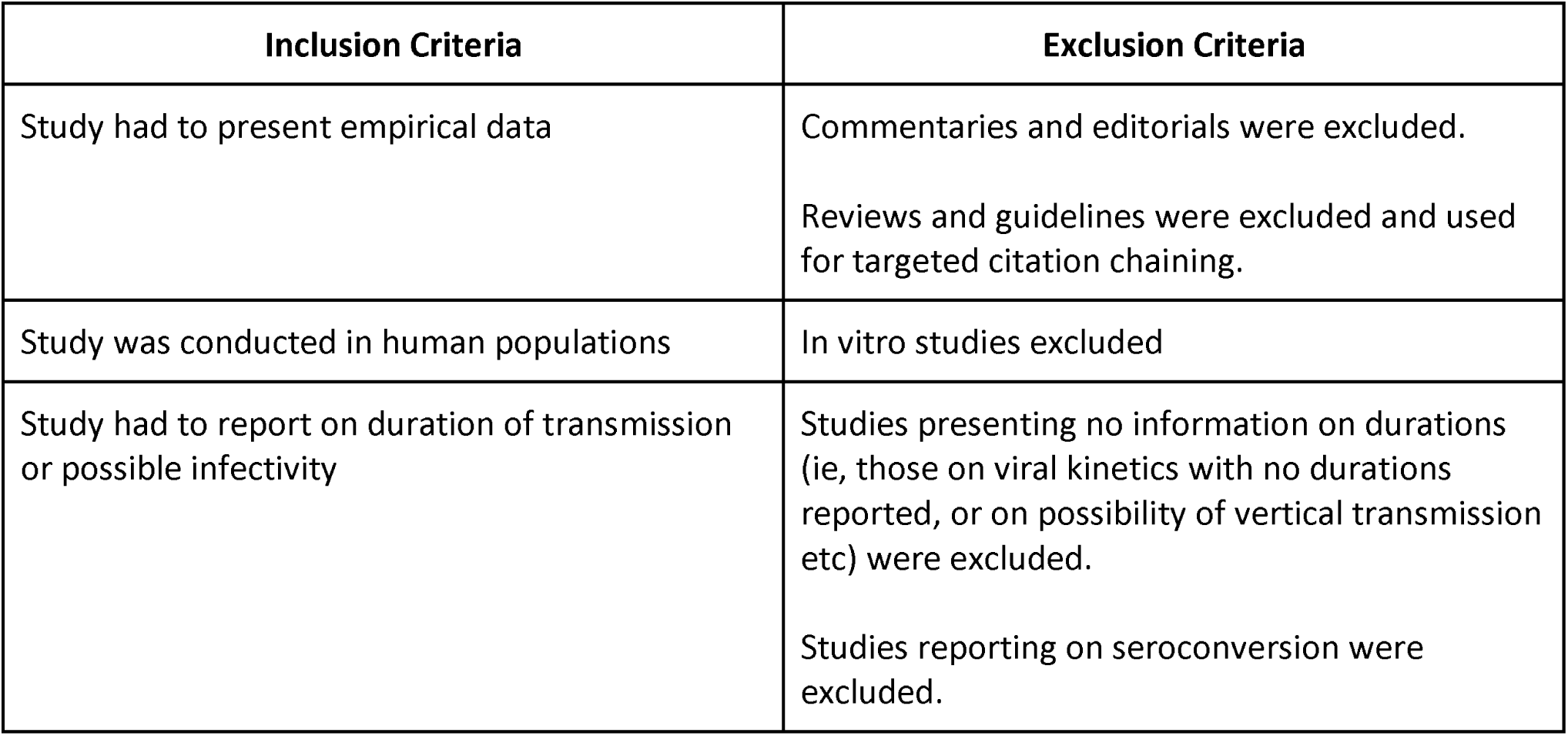

Appendix B – Inclusion/Exclusion Criteria:
| Inclusion Criteria | Exclusion Criteria |
| --- | --- |
| Study had to present empirical data | Commentaries and editorials were excluded.<br>Reviews and guidelines were excluded and used for targeted citation chaining. |
| Study was conducted in human populations | In vitro studies excluded |
| Study had to report on duration of transmission or possible infectivity | Studies presenting no information on durations (ie, those on viral kinetics with no durations reported, or on possibility of vertical transmission etc) were excluded.<br>Studies reporting on seroconversion were excluded. |

## SUPPLEMENTARY FIGURES

**Supplementary Figure 1.**
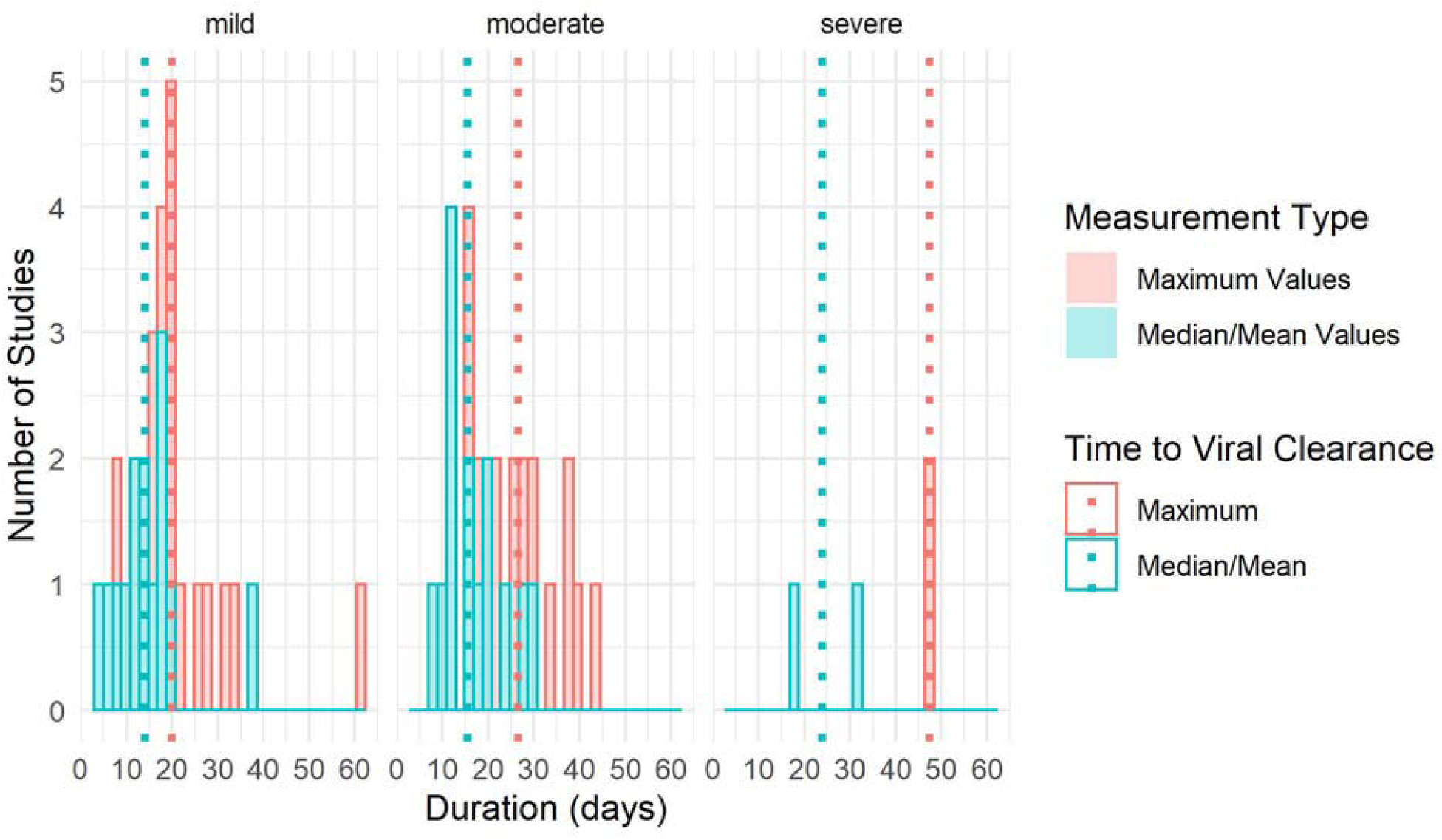
Histograms displaying the distribution of durations of communicability from studies reporting data on both median/mean and maximum durations to viral clearance, by disease severity (n=30). Data on median/mean values are shown in teal and maximum values are shown in pink. Dotted vertical lines represent median values across studies for median/mean duration (teal) and maximum duration (pink) to viral clearance.

## Notes

### Competing Interest Statement

The authors have declared no competing interest.

### Funding Statement

This study was not externally funded.

### Author Declarations

Ethics approval was not needed for this paper as it was a knowledge synthesis and did not collect patient-level data.

